# Novel Diffuse White Matter Abnormality Biomarker at Term-Equivalent Age Enhances Prediction of Long-term Motor Development in Children Born Very Preterm

**DOI:** 10.1101/2020.08.04.20167031

**Authors:** Nehal A. Parikh, Karen Harpster, Lili He, Venkata Sita Priyanka Illapani, Fatima Chughtai Khalid, Mark A. Klebanoff, T. Michael O’Shea, Mekibib Altaye

## Abstract

Our objective was to evaluate the independent prognostic value of a novel MRI biomarker – objectively diagnosed diffuse white matter abnormality volume (DWMA; diffuse excessive high signal intensity) – for prediction of motor outcomes in very preterm infants. We prospectively enrolled a geographically-based cohort of very preterm infants without severe brain injury and born before 32 weeks gestational age. Structural brain MRI was obtained at term-equivalent age and DWMA volume was objectively quantified using a published validated algorithm. These results were compared with visually classified DWMA. We used multivariable linear regression to assess the value of DWMA volume, independent of known predictors, to predict motor development as assessed using the Bayley Scales of Infant & Toddler Development, Third Edition at 3 years of age. The mean (SD) gestational age of the cohort was 28.3 (2.4) weeks. In multivariable analyses, controlling for gestational age, sex, and abnormality on structural MRI,DWMA volume was an independent prognostic biomarker of Bayley Motor scores (*β*= –12.59 [95% CI: –18.70, –6.48] R^2^=0.41). Conversely, visually classified DWMA was not predictive of motor development. In conclusion, objectively quantified DWMA is an independent prognostic biomarker of long-term motor development in very preterm infants and warrants further study.

---

Cerebral palsy (CP) describes a spectrum of life-long disorders of movement and posture that impacts 800,000 Americans^1^. CP is the most common physical disability in children, with annual healthcare costs of $15 billion^2^. Up to 10% of very preterm infants develop CP and 32-42% develop minor motor abnormalities^3,4,5^. Despite our understanding that these motor abnormalities are the result of abnormal development or brain injury during the fetal or neonatal period, children typically do not receive a diagnosis until 1 to 2 years of age^6^. There is wide consensus that earlier diagnosis, soon after birth, is urgently needed to take full advantage of critical windows of early neuroplasticity, particularly during the first two years^7,8^. Earlier diagnosis would facilitate targeted delivery of early interventions^9^ and novel habilitative therapies during this optimal period for brain development^10^.

Structural MRI (sMRI) at term-equivalent age is normal in up to 30% of diagnosed CP cases^11,12,13,14^. New advanced quantitative MRI measures may hold the greatest promise for enhancing prediction accuracy^15^ with quantitative cerebral morphometric analyses representing the most clinically feasible approach. Of these morphometric measures, objective assessment of diffuse excessive high signal intensity (DEHSI) abnormality is of import due to its high prevalence in preterm infants^14,16,17,18,19^, association with in-vivo microstructural^18,20^, metabolic^21^, and postmortem pathology^22^, and early evidence suggesting a correlation with neurodevelopmental impairments (NDI)^17,23,24,25,26^. While most studies that diagnosed DEHSI visually/qualitatively have not reported a significant association with NDI^18,19,27,28,29,30,31,32^, when quantified objectively, DEHSI appears to significantly predict cognitive and language development in extremely preterm infants^24,25^. To better reflect its pathologic nature, we will henceforth use the label diffuse white matter abnormality [DWMA] in place of DEHSI. The goal of this study was to examine the prognostic value of objectively quantified DWMA volume at term-equivalent age for prediction of motor development in a prospective cohort of very preterm infants. We hypothesized that objectively quantified DWMA volume would be an independent predictor of motor development at 3 years of age.

## Methods

### Population

All very preterm infants born at 31 weeks completed gestation or earlier and admitted to any of the four level III neonatal intensive care units (NICUs) in Columbus, Ohio from November 2014 to March 2016 were eligible for inclusion[25]. We prospectively enrolled 110 very preterm infants from a consecutively eligible sample of infants during this period. The four NICUs were Nationwide Children’s Hospital (NCH), Ohio State University Medical Center, Riverside Hospital, and Mount Carmel St. Ann’s Hospital. These NICUs care for approximately 80% of all very preterm infants in the Columbus, Ohio region. We excluded any infants with congenital or chromosomal anomalies that affected the central nervous system and likely result in a poor outcome. Data collection occurred between January 2015 and July 2018. The NCH Institutional Review Board approved the study at NCH and the other study sites through established reciprocity agreements. Written informed consent was obtained from a parent or guardian of each very preterm infant after they were given sufficient time to determine if they wished to participate. All methods/research activities were carried out in accordance with the NCH Institutional Review Board guidelines and regulations. All study infants were invited for routine developmental follow-up in the NCH High-Risk Follow-up Clinic up to 3 years corrected age.

### Magnetic Resonance Imaging Acquisition

We performed brain structural MRI scans on all 110 study infants at NCH on a 3T Siemens Skyra MRI scanner with at 32-channel pediatric head coil between the ages of 39 and 44 weeks post-menstrual age (PMA). Most infants from NCH were typically imaged while they were still inpatients, while all infants cared for at the other three NICUs were imaged as outpatients after being discharged. All inpatient MRI scans were attended by a skilled neonatal nurse and a neonatologist. Heart rate and oxygen saturation of all infants were monitored continuously during all scans. We performed all imaging without sedation by feeding the infants 30 minutes prior to the scan, applying silicone earplugs and swaddling the infants in a blanket and a vacuum immobilization device (MedVac, CFI Medical Solutions, Fenton, MI) to promote natural sleep. There were no adverse events. The following structural MRI sequence parameters were used for all infants: axial T2-weighted: echo time 147, repetition time 9500 ms, echo train length 16, flip angle 150°, resolution 0.93 x 0.93 x 1.0 mm^3^, scan time 4:09 min.; axial SWI: echo time 20,repetition time 27 ms, flip angle 15°, resolution 0.7 x 0.7 x 1.6 mm^3^, time 3:11 min.; 3-dimensional magnetization-prepared rapid gradient echo: echo time 2.9, repetition time 2270 ms, inversion recovery time 1600 ms, echo spacing time 8.5 ms, flip angle 13°, resolution 1.0 x 1.0 x 1.0 mm^3^, time 3:32 min. All infants tolerated the MRI procedure without adverse events.

### Image Post-Processing

We applied our previously published algorithm to objectively detect and quantify DWMA on T2-weighted MRI (Figure 1; For in-depth methods and additional examples of DWMA segmentation see He et al.)^25^. To summarize, first we conducted bias field correction (removal of signal intensity inhomogeneity caused mainly by the radiofrequency coils) and intensity normalization (reducing the variations in signal intensity and contrast across slices and across subjects). Next, we conducted brain tissue segmentation using a neonatal probabilistic brain atlas as a guide and defined DWMA to be any voxels with signal intensity values greater than ∝ standard deviations above the mean for all cerebral tissues (white and gray matter). We refer to ∝ as our cut-off threshold. For this study, we examined a cut-off thresholds of 2.0. However, this threshold was too restrictive and defined only very small regions as DWMA; therefore, we chose a lower threshold of 1.8 SD. We controlled for partial volume artifacts by only labeling voxels with high gray and white matter membership probability (≥95%) as cerebral tissues. We manually removed the few isolated false positive voxels detected by the algorithm. Total DWMA volume was calculated as the product of a single voxel volume (determined by the imaging resolution) and the total number of voxels in the detected DWMA region. We limited DWMA detection to the centrum semiovale only because we have found this to be the most predictive white matter region and it is not confounded by the normal high signal intensity of the periventricular crossroads^24,25,26^. We defined the centrum semiovale as the central white matter in the two slices immediately above the lateral ventricles on axial view. We calculated a normalized DWMA volume by dividing DWMA volume by total cerebral white matter volume. All analyses were performed masked to clinical and outcome data.

**Figure 1.**
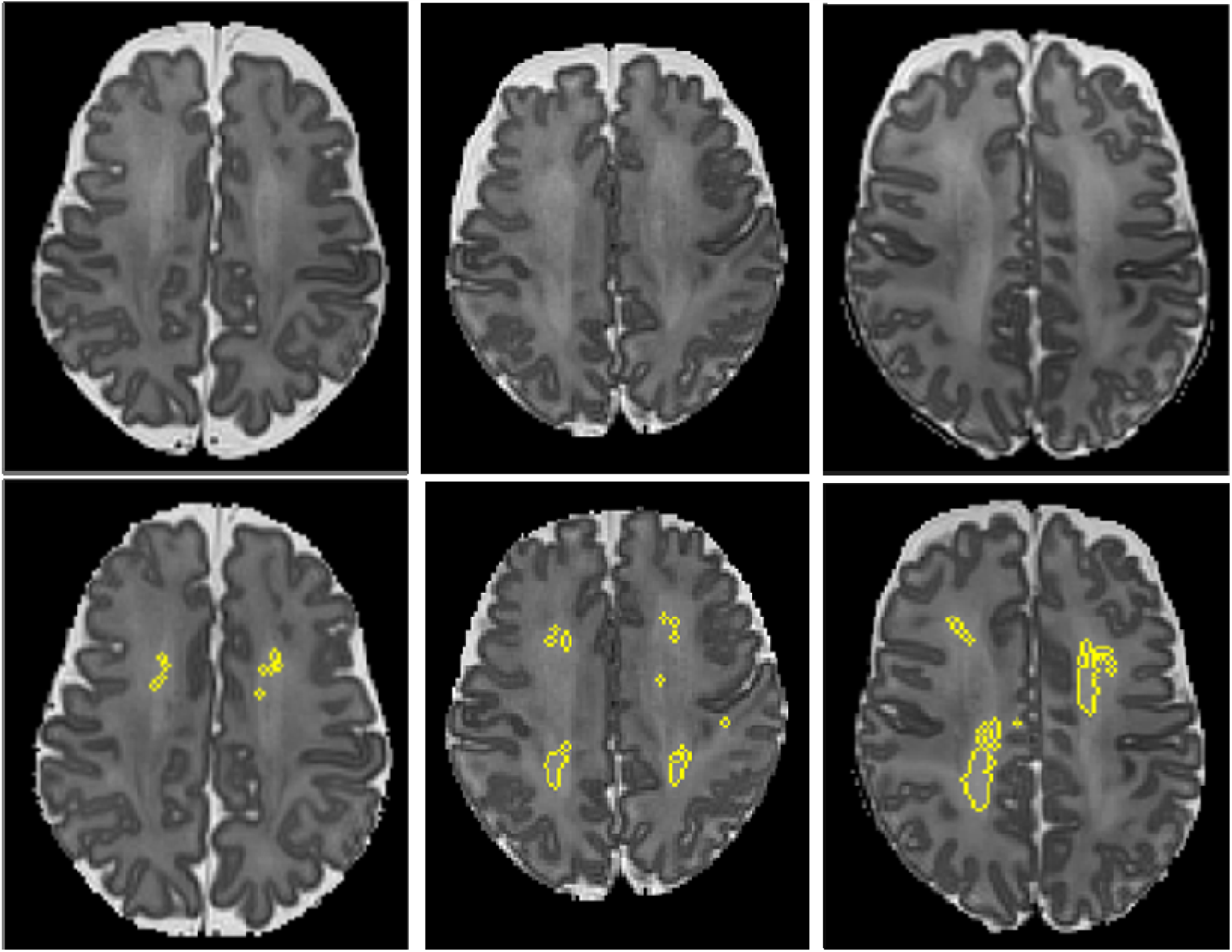
Objective segmentation of diffuse white matter abnormality (DWMA) in the centrum semiovale. The top three panels display raw axial T2-weighed MRI images through the centrum semiovale (immediately above the lateral ventricles) from very preterm infants born at 27 weeks (left), 26 weeks (center) and 31 weeks (right) gestation and imaged at term-equivalent age. Higher signal intensity than the subcortical white matter can be seen in the central white matter of the centrum semiovale, particularly for the 31-week gestation infant. The bottom panels display the corresponding slices with objectively segmented DWMA in yellow. The 27-week infant (left) was diagnosed with mild DWMA, the 26-week infant (center) was diagnosed with moderate DWMA, and the 31-week infant had severe DWMA.

### MRI Imaging Assessment

All brain structural MRI readings were performed by pediatric neuroradiologists who used a standardized scoring system graded for degree of brain injury/maturation, and the objective quantitative biometric measurements were performed separately by a trained expert, per Kidokoro et al.^33^. This approached yielded a global brain abnormality score, which was categorized as normal (total score, 0–3), mild (total score, 4–7), moderate (total score, 8–11), or severe abnormality (total score ≥12).

A single reader (NAP) with greater than 10 years of experience interpreting neonatal MRI scans performed visual qualitative classification of DWMA, masked to clinical and outcome data. The DWMA score was based on severity and extent as described by Kidokoro et al.^18^. Infants were assigned grade 0 if there was no DWMA or if high signal intensity was present only in the periventricular crossroads, grade 1 if DMWA was only visible in one region, grade 2 if DWMA was visible in two regions, and grade 3 if three or more regions were involved in addition to the normal signal intensity observed in the crossroads. While DWMA was observed in all white matter regions, the centrum semiovale was the most commonly identified region with qualitatively defined DWMA. The reader also assessed whether the margins of the posterior crossroads were invisible (defined as invisible posterior crossroads). The same reader reevaluated 20 randomly chosen MRI scans three weeks later and used kappa (κ) statistics to assess intra-rater agreement for DWMA grade. Of the 20 subjects, complete agreement was seen in 60% of cases (expected agreement 31.0%) for a κ of 0.42. This represents a fair to moderate agreement strength^34,35^.

### Neurodevelopmental Assessment

Participating infants underwent a comprehensive neurodevelopmental evaluation at a median age of 36.1 (IQR: 35.3-37.5) months in the NCH High-Risk Follow-up Clinic. We assessed overall motor development using the standardized Bayley Scales of Infant and Toddler Development,Third Edition (Bayley-III). A Motor composite score (composite of fine and gross motor development scores) that was 3 SD below the normative mean was assigned to children who could not complete the test due to difficulty resulting from likely severe disability. The composite score for the Bayley-III Motor subscale is scaled to metric with a mean of 100 (SD 15) and range of 40-160. Examiners performed the standardized Amiel-Tison neurologic exam,^36^ which included evaluation of tone, reflexes, posture, and strength; gross motor function was classified using the Gross Motor Function Classification System^37^. Cerebral palsy was defined a priori as abnormal muscle tone in at least one extremity and abnormal control of movement and posture. All assessments were performed by assessors who were masked to the quantitative DWMA diagnosis but not masked to clinical information.

### Statistical Analyses

In univariate analyses, we examined the relationship between the normalized DWMA volume and the Bayley-III Motor composite score using linear regression. To evaluate the independent prognostic value of DWMA volume, we performed multivariable regression by adding known perinatal predictors of Bayley score, including sex, gestational age, and global brain abnormality score. In addition, we also added center/NICU and PMA at MRI to the multivariable model to control for their potential confounding effects. We also tested a model that substituted global brain abnormality with a composite variable that included sMRI injury variables known to be strong predictors of motor impairment: cystic white matter abnormalities, hemorrhage (intraventricular, parenchymal, and/or cerebellar), and punctate white matter lesions^7^. The internal validity of our final model was tested by estimating a bias-corrected confidence interval derived from a bootstrap procedure involving 10,000 resamples^38^. In secondary analyses, to assess prediction accuracy for CP, we used Fisher’s exact test to evaluate prognostic properties, including sensitivity, specificity, positive and negative likelihood ratios, for the following three predictors (all defined a priori): 1) objectively diagnosed severe DWMA (normalized DWMA volume dichotomized at >90th percentile), 2) global brain abnormality (moderate or greater), and visually-classified severe DWMA (grade 3). We used logistic regression to evaluate the relationship between DWMA and CP. Last, we used Pearson’s correlation and multivariable linear regression to assess the relationship between 1) DWMA volume and global brain abnormality score and 2) DWMA volume and visually defined DWMA. We used the traditional two-sided *P* value of <0.05 to indicate statistical significance. All analyses were performed using STATA 16.0 (Stata Corp., College Station, TX).

## Results

Of the original cohort of 110 very preterm infants, we excluded one infant due to excessive motion artifacts and excluded all 11 infants with severe brain injury since this interfered with accurate DWMA segmentation (e.g. severe ventriculomegaly resulting in loss of centrum semiovale white matter). Structural MRI was performed at a mean (SD) PMA of 40.3 (0.5) weeks. By 3 years of age, 77 infants (79%) returned for Bayley motor testing and 82 of the 98 infants (84%) returned for cerebral palsy testing (Supplementary Figure 1). The baseline characteristics for infants who returned for follow-up were not significantly different from those who did not (Table 1). The mean (SD) Bayley-III raw Gross Motor, raw Fine Motor, and Composite Motor scores were 60.1 (5.6), 44.0 (5.0), and 90.8 (12.2), respectively. Cerebral palsy was diagnosed in six infants (7.3%). Four infants were diagnosed with spastic diplegia, one had spastic left hemiplegia, and one had spastic quadriplegia. The latter infant was classified as GMFCS level 4 while the other five infants were classified as GMFCS level 1.

**Table 1.** Baseline characteristics of very preterm infants with neurodevelopmental follow-up by 3 years of age and those without follow-up.

| **Clinical Variables** | **Infants with**<br>**Follow-up**<br>**(N=77)** | **Infants without**<br>**Follow-up**<br>**(N=21)** | ***P*** |
| --- | --- | --- | --- |
| Antenatal steroids (complete course within 7 days), N (%) | 39 (50.6%) | 7 (33.3%) | 0.219 |
| Multiple births, N (%) | 24 (31.2%) | 9 (42.9%) | 0.435 |
| Lower socioeconomic status, N (%) | 35 (45.5%) | 5 (23.8%) | 0.085 |
| Male, N (%) | 42 (54.5%) | 11 (52.4%) | 1.00 |
| Gestational age at birth (weeks), mean (SD) | 28.3 (2.4) | 28.6 (3.0) | 0.596 |
| Birth weight (grams), mean (SD) | 1126 (396) | 1146 (407) | 0.838 |
| Transitional hypotension, N (%) | 6 (7.8%) | 2 (9.5%) | 1.00 |
| Sepsis (culture positive), N (%) | 9 (11.7%) | 6 (28.6%) | 0.084 |
| Postnatal steroids for bronchopulmonary dysplasia (BPD), N (%) | 6 (7.8%) | 0 | 0.336 |
| BPD (O<sub>2</sub> supplementation at 36 weeks postmenstrual age), N (%) | 38 (49.4%) | 10 (47.6%) | 1.00 |
| Global brain injury score, median (IQR) | 3 (1, 4) | 2 (1, 3) | 0.319 |
| Normalized DWMA volume, median (IQR) | .0073 (.0006, .0264) | .0054 (.0006, .0268) | 0.698 |

Based on the global brain abnormality score, five infants were classified as having moderate injury (6.5%), 19 had mild injury (24.7%), and 53 had no injury (68.8%) on their sMRI at term-equivalent age. Moderate injury was noted on sMRI in three of the six infants (50%) with CP. As stated above, all infants with severe injury were excluded from the study. Visually/qualitatively classified DWMA was diagnosed as severe (grade 3) in 11 infants (13.4%), moderate (grade 2) in 22 infants (26.8%), and no/mild (grade 0/1) in 49 infants (59.8%). Only one infant was diagnosed with invisible posterior crossroads.

In univariate analyses, DWMA volume was significantly predictive of Bayley-III Motor scores, explaining 26% of the variance in motor development (Table 2; Figure 2). This association remained significant even when raw DWMA volume was tested in the regression analyses, suggesting that the normalization by total white matter volume did not have a significant effect on the association with Bayley Motor scores. In multivariable analyses, controlling for other known predictors of Bayley scores, including sex, gestational age, and global brain abnormality, normalized DWMA volume (*β*=–12.59 [95% CI: –18.70, –6.48]) remained a significant predictor of Bayley Motor development at age 3 (Table 2). Replacing global brain abnormality score with cystic abnormalities, hemorrhage, and punctate white matter lesion variables actually reduced the model adjusted R^2^ (38.9%) and enhanced the predictive power of DWMA (*β*=– 14.33). The bootstrap bias-corrected confidence intervals were comparable (*β* 95% CI: –18.60, – 4.31), supporting the internal validity of the final model.

**Table 2.**
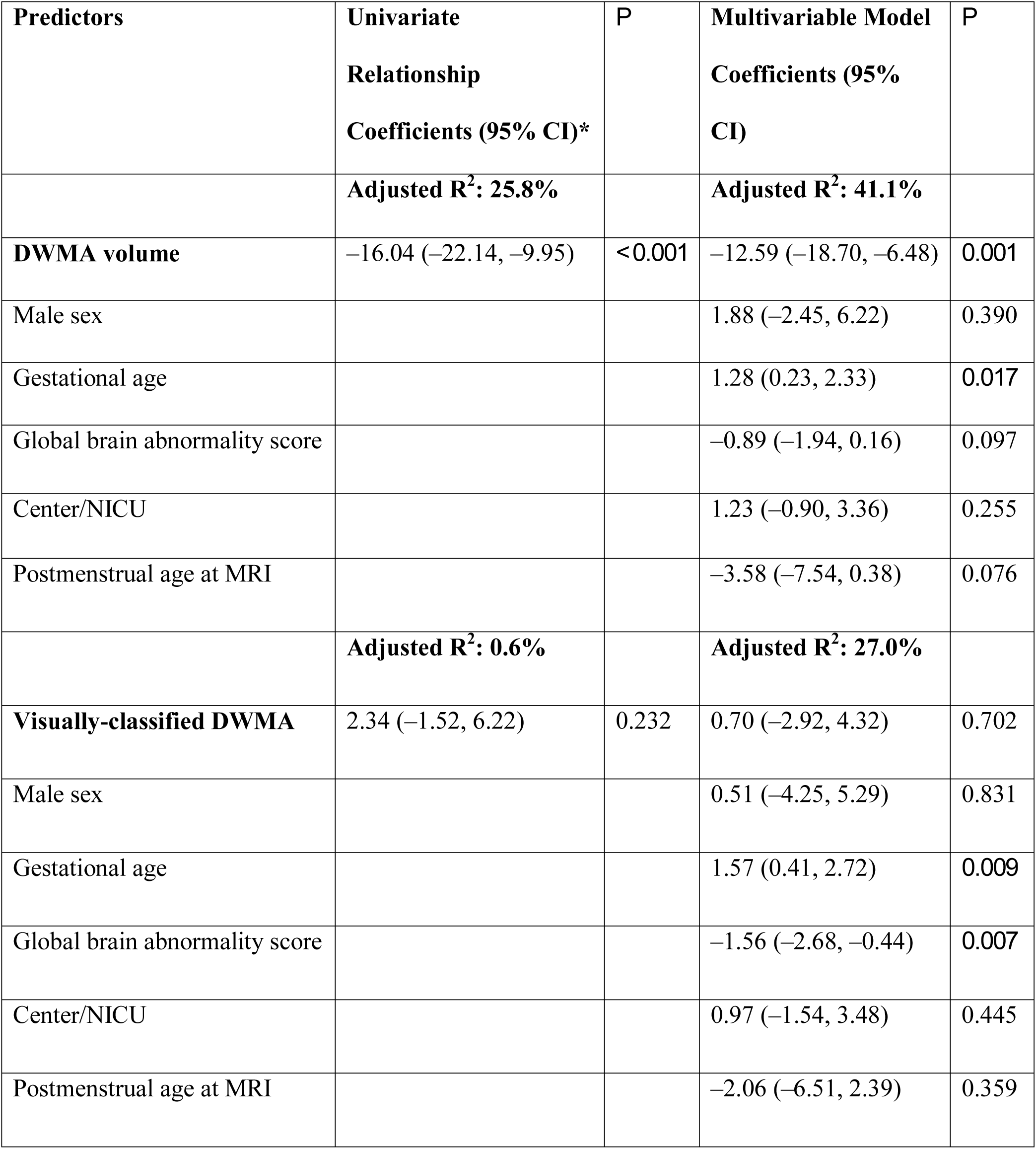
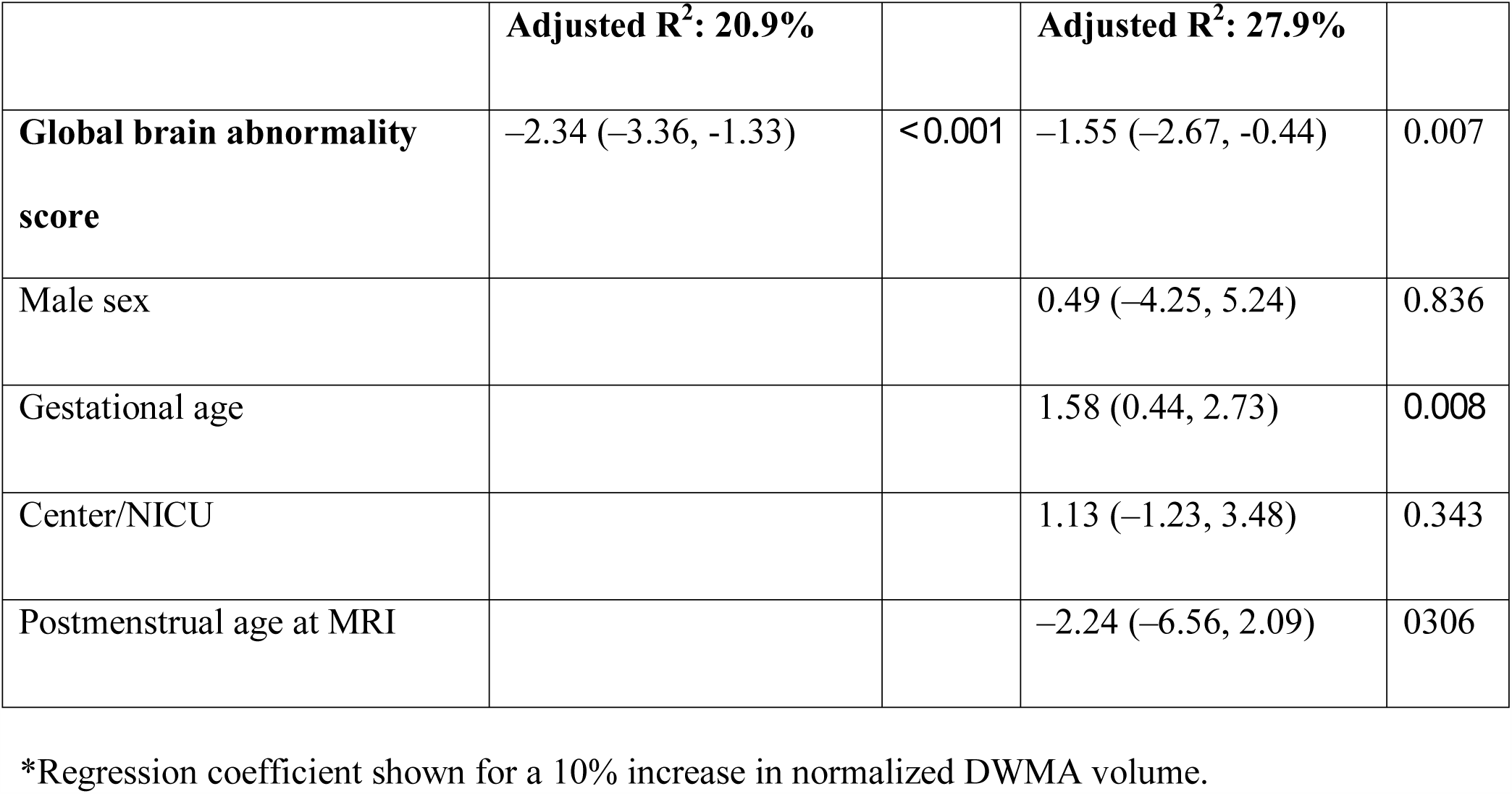
Regression coefficients for linear regression models of objectively quantified, normalized DWMA volume versus visually-classified DWMA as predictors of Bayley-III Motor composite score in very preterm infants.

| **Predictors** | **Univariate Relationship Coefficients (95% CI)\*** | ***P*** | **Multivariable Model Coefficients (95% CI)** | ***P*** |
| --- | --- | --- | --- | --- |
|  | **Adjusted R<sup>2</sup>: 25.8%** |  | **Adjusted R<sup>2</sup>: 41.1%** |  |
| **DWMA volume** | -16.04 (-22.14, -9.95) | <0.001 | -12.59 (-18.70, -6.48) | 0.001 |
| Male sex |  |  | 1.88 (-2.45, 6.22) | 0.390 |
| Gestational age |  |  | 1.28 (0.23, 2.33) | 0.017 |
| Global brain abnormality score |  |  | -0.89 (-1.94, 0.16) | 0.097 |
| Center/NICU |  |  | 1.23 (-0.90, 3.36) | 0.255 |
| Postmenstrual age at MRI |  |  | -3.58 (-7.54, 0.38) | 0.076 |
|  | **Adjusted R<sup>2</sup>: 0.6%** |  | **Adjusted R<sup>2</sup>: 27.0%** |  |
| **Visually-classified DWMA** | 2.34 (-1.52, 6.22) | 0.232 | 0.70 (-2.92, 4.32) | 0.702 |
| Male sex |  |  | 0.51 (-4.25, 5.29) | 0.831 |
| Gestational age |  |  | 1.57 (0.41, 2.72) | 0.009 |
| Global brain abnormality score |  |  | -1.56 (-2.68, -0.44) | 0.007 |
| Center/NICU |  |  | 0.97 (-1.54, 3.48) | 0.445 |
| Postmenstrual age at MRI |  |  | -2.06 (-6.51, 2.39) | 0.359 |

|  | **Adjusted R<sup>2</sup>: 20.9%** |  | **Adjusted R<sup>2</sup>: 27.9%** |  |
| --- | --- | --- | --- | --- |
| **Global brain abnormality score** | -2.34 (-3.36, -1.33) | *<0.001* | -1.55 (-2.67, -0.44) | 0.007 |
| Male sex |  |  | 0.49 (-4.25, 5.24) | 0.836 |
| Gestational age |  |  | 1.58 (0.44, 2.73) | 0.008 |
| Center/NICU |  |  | 1.13 (-1.23, 3.48) | 0.343 |
| Postmenstrual age at MRI |  |  | -2.24 (-6.56, 2.09) | 0.306 |

\*Regression coefficient shown for a 10% increase in normalized DWMA volume.

**Figure 2.**
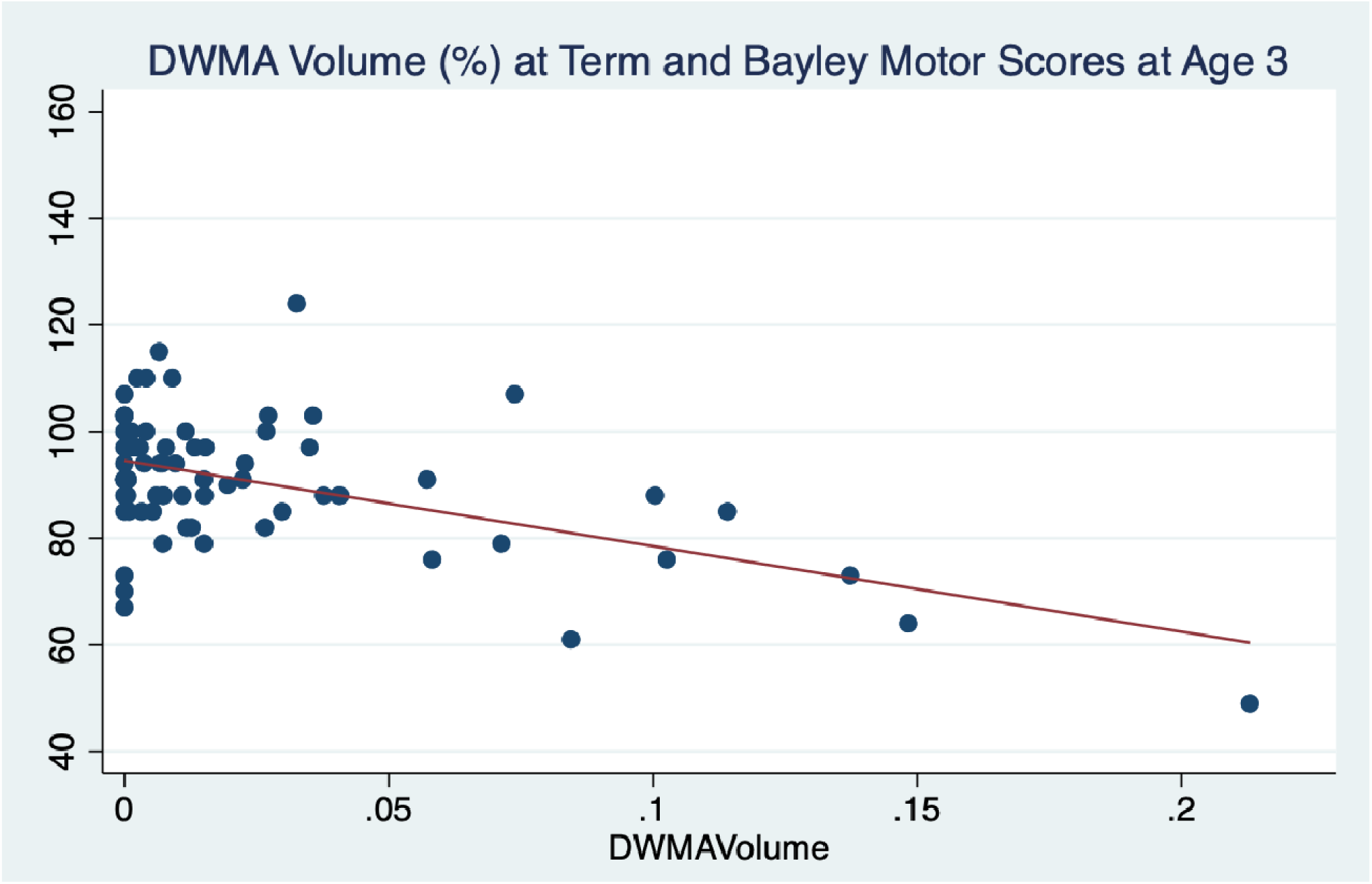
Scatterplot demonstrating relationship between objectively quantified normalized DWMA volume (%) at term-equivalent age and observed Bayley Scales Motor composite scores at 3 years of age.

To confirm that the significant predictive relationship we observed between normalized volume of DWMA and Motor scores (t= –4.11; p<0.001) was not a function of white matter volume loss, we replaced normalized DWMA volume with DWMA volume that was uncorrected for white matter volume. This replacement did not result in a meaningful difference in the model parameters (t=–3.92; p<0.001). When we control for the effects of differencing head sizes by including total white matter in the multivariable model (with uncorrected DWMA volume), the model remains very comparable (t=–3.89; p<0.001); when controlled for total intracranial volume, the results again remain comparable (t=–3.95; p<0.001). The total explained variance in Bayley Motor scores was 39% and 40%, respectively (compared to 41% for normalized DWMA volume). These analyses suggest that DWMA is independent of other white matter pathology.

Visual, qualitative diagnosis of DWMA was not significantly predictive of Motor scores in univariate analyses (P=.23). Inclusion of known predictors and confounders in the model did not substantially change this relationship (P=.70; Table 2). Global brain abnormality score was also a significant predictor of Motor scores, even after controlling for other predictors. However, it was not as predictive as DWMA volume. The addition of DWMA volume increased the variance in Bayley Motor scores by another 13.2% (41.1% vs. 27.9%; Table 2). Finally, normalized DWMA volume was significantly correlated with global brain abnormality score (r=0.30; p=0.003) but not with visually defined qualitative DWMA (r=0.09; p=0.23) in univariate analyses. In multivariable linear regression models, controlling for gestational age, sex, PMA at MRI, and center, we observed a significant relationship between objectively defined DWMA volume and global brain abnormality score (*β*=0.032; 95% CI: 0.003, 0.062; p=0.032). In similarly controlled multivariable linear regression models, we did not observe a significant relationship between objectively defined DWMA volume and visually defined DWMA (*β*=0.044; 95% CI: - 0.064, 0.152; p=0.418).

In secondary logistic regression analyses, a 10% increase in DWMA volume was associated with an odds ratio of 31.64 (95% CI: 3.96, 253.03) of developing CP (p<0.001). A one point increase in global brain abnormality was associated with an odds ratio of 2.25 (95% CI: 1.44, 3.51) of developing CP (p<0.001). The relationship between DWMA volume and CP remained significant in a multivariable model after controlling for global brain abnormality, gestational age, and center (OR 12.64, 95% CI: 1.41, 113.43). Conversely, we did not find a significant relationship between qualitatively diagnosed DWMA and CP (OR 2.29; 95% CI: 0.81, 6.46; p=0.118). Objectively quantified severe DWMA (P<0.001) and global brain abnormality on structural MRI (P=0.004) were both significantly predictive of CP, while visually-classified severe DWMA (grade 3) was a poor predictor of CP (P=1.00) (Table 3).

**Table 3.** Prognostic test properties for objectively-diagnosed severe DWMA, abnormal structural MRI, and visually-classified severe diffuse white matter abnormality (DWMA) for predicting cerebral palsy in very preterm infants.

| **Predictors** | **Sensitivity**<br>**(95% CI)** | **Specificity**<br>**(95% CI)** | **Positive**<br>**Likelihood**<br>**Ratio**<br>**(95% CI)** | **Negative**<br>**Likelihood**<br>**Ratio**<br>**(95% CI)** | ***P*** |
| --- | --- | --- | --- | --- | --- |
| Objectively diagnosed<br>severe DWMA | 66.7%<br>(22.3%, 95.7%) | 94.7%<br>(87.1%, 98.6%) | 12.7<br>(4.2, 38.4) | 0.35<br>(0.11, 1.09) | *<0.001* |
| Global brain<br>abnormality | 50.0%<br>(11.8%, 88.2%) | 96.1%<br>(88.9%, 99.2%) | 12.7<br>(3.2, 49.7) | 0.52<br>(0.23, 1.16) | *0.004* |
| Visually-classified<br>severe DWMA | 16.7%<br>(0.42%, 64.1%) | 86.8%<br>(77.1%, 93.5%) | 1.27<br>(0.2, 8.3) | 0.96<br>(0.66, 1.39) | 1.00 |

## Discussion

We demonstrated that objectively quantified DWMA is a significant and independent prognostic biomarker of motor development at 3 years of age in very preterm infants. Normalized DWMA volume remained a prominent predictor of standardized Motor scores even after controlling for other known predictors of motor development such as sMRI abnormalities, gestational age, and sex. This is notable because we excluded all infants with severe brain injury, which is the most prominent risk factor for the development of CP and motor impairments. Of the six infants that developed CP in this cohort, five had mild CP. Two of the three infants with a normal sMRI developed spastic diplegia. A recent CP registry study found that infants with these CP subtypes were twice as likely to have normal sMRI scans^39^. Our results suggest that DWMA is pathologic and deserves further testing in larger studies to externally validate its prognostic value for the early detection of minor motor impairments and CP.

In two previous independent cohorts, we have shown that DWMA volume quantified using our objective algorithm is predictive of cognitive and language development at 2 years^25,26^. For motor development, it is difficult to compare our objective DWMA biomarker results with other studies because no prior study has attempted to predict Motor outcomes using objectively quantified DWMA. However, similar to our findings, multiple studies have examined the link between visually classified DWMA and motor development and found no correlation^17,18,19,23,27,28,29,30,32^. There are likely several confounding factors that contributed to a lack of association. First, visually diagnosed DWMA is subjective and exhibits suboptimal retest reliability, as demonstrated in our study and several prior studies^14,31,40,41^. This may be due to the signal inhomogeneity and the occurrence of developmental crossroads that are routinely present on all MRI scans at term-equivalent age. This is especially true for periventricular white matter regions, which potentially explains why our previous study showed lower predictive value for this white matter region as compared to the centrum semiovale[24]. Therefore, we limited assessment of DWMA to only the centrum semiovale white matter. Lower diagnostic reliability increases measurement error, which can reduce the likelihood of finding a significant association^42^. Our intra-rater reliability for visual diagnosis of DWMA was only fair to moderate (K 0.42). This reliability was comparable to our prior study where a pediatric neuroradiologist diagnosed DWMA (K 0.46)^14^. Second, for a given sample size, a qualitative diagnosis is categorical and will therefore exhibit lower study power than a quantitative diagnosis (continuous measure)^43,44^. Lastly, even when qualitative DWMA diagnostic reliability is excellent^18^, DWMA diagnosis may still be inaccurate because there is no gold standard test to confirm true DWMA pathology.

Our DWMA segmentation algorithm is automated, easy to use, and can generate DWMA and whole-brain tissue volumes within 5 minutes. For about half of the MRI scans, it requires no further manual correction and the other half it incorrectly labels between 2 to 8 voxels, most commonly in the interhemispheric fissure or peripheral white matter. While we currently remove these mislabeled voxels manually, we have recently developed a fully-automated approach using machine learning to overcome this limitation^45^. Such a tool could be used to facilitate clinical translation, as it can be readily integrated into clinical MRI platforms to generate DWMA volumes immediately at the point of care, following sMRI acquisition at term-equivalent age.

At term-equivalent age, sMRI is the most accurate test for early detection of CP. Although it’s predictive accuracy has been touted as approximately 90%^7^, when more robust evidence is considered, its sensitivity is closer to 70% and its predictive probability is only 35%^11,12,13,14,46^. This leaves a substantial gap in our ability to accurately detect CP at term-equivalent age in order to take advantage of the early critical window for neuroplasticity in the first two years. This is the period during which proven (re)habilitative interventions could restore motor function and thus improve quality of life. Prognostic tests such as the general movements assessment and the Hammersmith Infant Neurological Exam are being increasingly performed in many centers at 3 months corrected age,^7^ however more research is still needed to determine their accuracy in combination with injury on sMRI in predicting CP and minor motor abnormalities^47^. Other advanced quantitative MRI measures such as diffusion MRI and structural and functional sensorimotor tract connectivity^48^ could potentially fill this gap, as highlighted in a recent systematic review^15^. For example, several studies have reported abnormal microstructural properties of DWMA using diffusion MRI^18,20^. However, when these measures were compared to quantitative DWMA volumes determined via signal intensity, the addition of microstructural measures did not improve outcome prediction^24^. The incremental predictive value of other promising advanced MRI biomarkers, independent of sMRI, remains to be validated in larger, population-based cohort studies.

Our study has several limitations. Our follow-up rate was 79%, which may have introduced ascertainment bias. However, the baseline characteristics of infants with and without follow-up testing were comparable, suggesting a low risk of bias. Only six infants developed CP and thus our secondary CP prognostic analyses will need to be validated in larger studies. Also, we were unable to determine the predictive ability of DWMA volume over and above general movements assessment or early standardized motor exam because these tests were not part of the research study or clinical care during study enrollment. This limitation is being addressed in our newer and larger cohort study. Our Bayley assessors were blinded to DWMA result but not blinded to clinical or structural MRI information; thus, this may have biased our findings. However, this bias would reduce the independent association of DWMA volume with Bayley Motor scores by potentially strengthening the association of clinical and global brain abnormality scores with Motor scores. Strengths of our study include a geographically-based cohort, objective quantification of DWMA on sMRI that can be readily translated clinically, comparison with the current standard method of visual classification, standardized assessments of motor outcomes up to 3 years of age when minor motor abnormalities are more evident, and independent validation of DWMA volume as a new prognostic biomarker, over and above existing predictors.

In conclusion, in this multicenter prospective cohort study, we were able to demonstrate for the first time that objectively quantified DWMA is an independent predictor of motor development in very preterm infants. We also validated prior research showing that visually classified DWMA is not predictive of neurodevelopmental outcomes and is therefore suboptimal for use in clinical practice. Additional studies are needed to externally validate the use of DWMA volume as an early prognostic biomarker for cerebral palsy and minor motor impairments and to enable clinical translation of our DWMA algorithm. If externally validated, our findings could be applied to improve risk stratification at hospital discharge for targeted, aggressive early intervention therapies.

## Data Availability

All data, software, and code from this study can be accessed from the lead author upon request

## Data sharing

All data, software, and code from this study are being submitted for publication and can be accessed from the lead author in the meantime.

## Acknowledgements

This study was supported by the National Institutes of Health (grants R01-NS094200 and R01-NS096037 [to NAP]; and grant R21-HD094085 and Trustee grant from Cincinnati Children’s Hospital Medical Center [to LH]). The funders played no role in the design, analysis, or presentation of the findings. We sincerely thank Jennifer Notestine, RN and Valerie Marburger, NNP for serving as the study coordinators, Josh Goldberg, MD for assisting with recruitment and Mark Smith, MS, for serving as the study MR technologist. We are also grateful to the families, NICU personnel, and High-Risk clinic staff that made this study possible.

## Author Contributions

N.A.P. conceived the experiments and wrote the manuscript; V.S.P.I. and L.H. conducted the experiments, N.A.P and M.A. performed the statistical analyses, N.A.P., M.K, and M.A. analyzed the results. T.M.O, K.H. and F.C.K provided critical feedback on the manuscript. All authors reviewed and provided critical feedback on the manuscript.

## Competing Interests

The authors declare no competing interests.

### Funding

Supported by the National Institutes of Health (grants R01-NS094200 and R01-NS096037 [to NAP] and grant R21-HD094085 [to LH]).

## Notes

### Competing Interest Statement

The authors have declared no competing interest.

### Author Declarations

The Nationwide Children's Hospital (NCH) Institutional Review Board approved the study at NCH and the other study sites through established reciprocity agreements.

## REFERENCES

1. Shevell, M. Cerebral palsy to cerebral palsy spectrum disorder: Time for a name change? Neurology, doi:10.1212/WNL.0000000000006747 (2018).

2. Honeycutt, A. A. et al. in Using Survey Data to Study Disability: Results From the National Health Interview Survey on Disability Vol. 3 (eds B.M. Altman, S.N. Barnartt, G.E. Hendershot, & S.A. Larson) 207–228 (Emerald Group Publishing Limited, 2003).

3. Sellier, E. et al.. Decreasing prevalence in cerebral palsy: a multi-site European population-based study, 1980 to 2003. Dev Med Child Neurol 58, 85–92, doi:10.1111/dmcn.12865 (2016).

4. Williams, J., Lee, K. J. & Anderson, P. J. Prevalence of motor-skill impairment in preterm children who do not develop cerebral palsy: a systematic review. Dev Med Child Neurol 52, 232–237, doi:10.1111/j.1469-8749.2009.03544.x (2010).

5. Van Hus, J. W., Potharst, E. S., Jeukens-Visser, M., Kok, J. H. & Van Wassenaer-Leemhuis, A. G. Motor impairment in very preterm-born children: links with other developmental deficits at 5 years of age. Dev Med Child Neurol 56, 587–594 (2014).

6. McIntyre, S., Morgan, C., Walker, K. & Novak, I. Cerebral palsy--don’t delay. Dev Disabil Res Rev 17, 114–129, doi:10.1002/ddrr.1106 (2011).

7. Novak, I. et al. Early, Accurate Diagnosis and Early Intervention in Cerebral Palsy: Advances in Diagnosis and Treatment. JAMA Pediatr 171, 897–907, doi:10.1001/jamapediatrics.2017.1689 (2017).

8. Johnston, M. V. Plasticity in the developing brain: implications for rehabilitation. Dev Disabil Res Rev 15, 94–101, doi:10.1002/ddrr.64 (2009).

9. Spittle, A., Orton, J., Anderson, P. J., Boyd, R. & Doyle, L. W. Early developmental intervention programmes provided post hospital discharge to prevent motor and cognitive impairment in preterm infants. Cochrane Database Syst Rev, CD005495, doi:10.1002/14651858.CD005495.pub4 (2015).

10. Morgan, C., Novak, I., Dale, R. C., Guzzetta, A. & Badawi, N. Single blind randomised controlled trial of GAME (Goals - Activity - Motor Enrichment) in infants at high risk of cerebral palsy. Res Dev Disabil 55, 256–267, doi:10.1016/j.ridd.2016.04.005 (2016).

11. Benini, R., Dagenais, L., Shevell, M. I. & Registre de la Paralysie Cerebrale au Quebec, C. Normal imaging in patients with cerebral palsy: what does it tell us? J Pediatr 162, 369–374 e361, doi:10.1016/j.jpeds.2012.07.044 (2013).

12. Van’t Hooft, J. et al. Predicting developmental outcomes in premature infants by term equivalent MRI: systematic review and meta-analysis. Syst Rev 4, 71, doi:10.1186/s13643-015-0058-7 (2015).

13. Hintz, S. R. et al. Neuroimaging and neurodevelopmental outcome in extremely preterm infants. Pediatrics 135, e32–42, doi:10.1542/peds.2014-0898 (2015).

14. Slaughter, L. A., Bonfante-Mejia, E., Hintz, S. R., Dvorchik, I. & Parikh, N. A. Early Conventional MRI for Prediction of Neurodevelopmental Impairment in Extremely-Low-Birth-Weight Infants. Neonatology 110, 47–54, doi:10.1159/000444179 (2016).

15. Parikh, N. A. Advanced neuroimaging and its role in predicting neurodevelopmental outcomes in very preterm infants. Semin Perinatol 40, 530–541, doi:10.1053/j.semperi.2016.09.005 (2016).

16. Maalouf, E. F. et al. Magnetic resonance imaging of the brain in a cohort of extremely preterm infants. J Pediatr 135, 351–357 (1999).

17. Dyet, L. E. et al. Natural history of brain lesions in extremely preterm infants studied with serial magnetic resonance imaging from birth and neurodevelopmental assessment. Pediatrics 118, 536–548, doi:10.1542/peds.2005-1866 (2006).

18. Kidokoro, H., Anderson, P. J., Doyle, L. W., Neil, J. J. & Inder, T. E. High signal intensity on T2-weighted MR imaging at term-equivalent age in preterm infants does not predict 2-year neurodevelopmental outcomes. AJNR Am J Neuroradiol 32, 2005–2010, doi:10.3174/ajnr.A2703 (2011).

19. Jeon, T. Y. et al. Neurodevelopmental outcomes in preterm infants: comparison of infants with and without diffuse excessive high signal intensity on MR images at near-term-equivalent age. Radiology 263, 518–526, doi:10.1148/radiol.12111615 (2012).

20. Counsell, S. J. et al. Axial and radial diffusivity in preterm infants who have diffuse white matter changes on magnetic resonance imaging at term-equivalent age. Pediatrics 117, 376–386, doi:10.1542/peds.2005-0820 (2006).

21. Wisnowski, J. L. et al. Altered glutamatergic metabolism associated with punctate white matter lesions in preterm infants. PLoS One 8, e56880, doi:10.1371/journal.pone.0056880 (2013).

22. Parikh, N. A., Pierson, C. R. & Rusin, J. A. Neuropathology Associated With Diffuse Excessive High Signal Intensity Abnormalities on Magnetic Resonance Imaging in Very Preterm Infants. Pediatr Neurol 65, 78–85, doi:10.1016/j.pediatrneurol.2016.07.006 (2016).

23. Iwata, S. et al. Qualitative brain MRI at term and cognitive outcomes at 9 years after very preterm birth. Pediatrics 129, e1138–1147, doi:10.1542/peds.2011-1735 (2012).

24. Parikh, N. A. et al. Automatically quantified diffuse excessive high signal intensity on MRI predicts cognitive development in preterm infants. Pediatr Neurol 49, 424–430, doi:10.1016/j.pediatrneurol.2013.08.026 (2013).

25. He, L. & Parikh, N. A. Atlas-guided quantification of white matter signal abnormalities on term-equivalent age MRI in very preterm infants: findings predict language and cognitive development at two years of age. PLoS One 8, e85475, doi:10.1371/journal.pone.0085475 (2013).

26. Parikh, N. A. et al. Objectively-Quantified Diffuse White Matter Abnormality at Term-Equivalen Age is an Independent Predictor of Neurodevelopmental Outcomes in very Preterm Infants. J Pediatr 220:56–63. doi: 10.1016/j.jpeds.2020.01.034 (2020).

27. Brostrom, L. et al. Clinical Implications of Diffuse Excessive High Signal Intensity (DEHSI) on Neonatal MRI in School Age Children Born Extremely Preterm. PLoS One 11, e0149578, doi:10.1371/journal.pone.0149578 (2016).

28. Hart, A. et al. Neuro-developmental outcome at 18 months in premature infants with diffuse excessive high signal intensity on MR imaging of the brain. Pediatr Radiol 41, 1284–1292, doi:10.1007/s00247-011-2155-7 (2011).

29. de Bruine, F. T. et al. Clinical implications of MR imaging findings in the white matter in very preterm infants: a 2-year follow-up study. Radiology 261, 899–906, doi:10.1148/radiol.11110797 (2011).

30. Skiold, B. et al. Neonatal magnetic resonance imaging and outcome at age 30 months in extremely preterm infants. J Pediatr 160, 559–566 e551, doi:10.1016/j.jpeds.2011.09.053 (2012).

31. Calloni, S. F. et al. Neurodevelopmental outcome at 36 months in very low birth weight premature infants with MR diffuse excessive high signal intensity (DEHSI) of cerebral white matter. Radiol Med 120, 1056–1063, doi:10.1007/s11547-015-0540-2 (2015).

32. Murner-Lavanchy, I. M. et al. Thirteen-Year Outcomes in Very Preterm Children Associated with Diffuse Excessive High Signal Intensity on Neonatal Magnetic Resonance Imaging. J Pediatr, doi:10.1016/j.jpeds.2018.10.016 (2018).

33. Kidokoro, H., Neil, J. J. & Inder, T. E. New MR imaging assessment tool to define brain abnormalities in very preterm infants at term. AJNR Am J Neuroradiol 34, 2208–2214, doi:10.3174/ajnr.A3521 (2013).

34. Landis, J. R. & Koch, G. G. The measurement of observer agreement for categorical data. Biometrics 33, 159–174 (1977).

35. McHugh, M. L. Interrater reliability: the kappa statistic. Biochem Med (Zagreb) 22, 276–282 (2012).

36. Amiel-Tison, C. & Gosselin, J. Neurological Development from Birth to Six Years., (The John Hopkins University Press, 1998).

37. Palisano, R. et al. Development and reliability of a system to classify gross motor function in children with cerebral palsy. Dev Med Child Neurol 39, 214–223 (1997).

38. Moons, K. G. et al. Risk prediction models: I. Development, internal validation, and assessing the incremental value of a new (bio)marker. Heart 98, 683–690, doi:10.1136/heartjnl-2011-301246 (2012).

39. Springer, A. et al. Profile of children with cerebral palsy spectrum disorder and a normal MRI study. Neurology, doi:10.1212/WNL.0000000000007726 (2019).

40. Morel, B., Antoni, G., Teglas, J. P., Bloch, I. & Adamsbaum, C. Neonatal brain MRI: how reliable is the radiologist’s eye? Neuroradiology 58, 189–193, doi:10.1007/s00234-015-1609-2 (2016).

41. Hart, A. R., Smith, M. F., Rigby, A. S., Wallis, L. I. & Whitby, E. H. Appearances of diffuse excessive high signal intensity (DEHSI) on MR imaging following preterm birth. Pediatr Radiol 40, 1390–1396, doi:10.1007/s00247-010-1633-7 (2010).

42. Coggon, D. in Epidemiology for the Uninitiated (ed BMJ) (BMJ Pub. Group, 1997).

43. Campbell, M. J., Julious, S. A. & Altman, D. G. Estimating sample sizes for binary, ordered categorical, and continuous outcomes in two group comparisons. BMJ 311, 1145–1148 (1995).

44. Altman, D. G. & Royston, P. The cost of dichotomising continuous variables. BMJ 332, 1080, doi:10.1136/bmj.332.7549.1080 (2006).

45. Li, H. et al. Objective and Automated Detection of Diffuse White Matter Abnormality in Preterm Infants Using Deep Convolutional Neural Networks. Front Neurosci 13, 610, doi:10.3389/fnins.2019.00610 (2019).

46. Nongena, P., Ederies, A., Azzopardi, D. V. & Edwards, A. D. Confidence in the prediction of neurodevelopmental outcome by cranial ultrasound and MRI in preterm infants. Arch Dis Child Fetal Neonatal Ed 95, F388–390, doi:10.1136/adc.2009.168997 (2010).

47. Parikh, N. A. Are Structural Magnetic Resonance Imaging and General Movements Assessment Sufficient for Early, Accurate Diagnosis of Cerebral Palsy? JAMA Pediatr 172, 198–199, doi:10.1001/jamapediatrics.2017.4812 (2018).

48. Parikh, N. A., Hershey, A. & Altaye, M. Early Detection of Cerebral Palsy Using Sensorimotor Tract Biomarkers in Very Preterm Infants. Pediatr Neurol, doi:10.1016/j.pediatrneurol.2019.05.001 (2019).

